# Lumbar Spine Degenerative Classification Using YOLO v8 and DeepScoreNet

**DOI:** 10.1101/2024.12.06.24318595

**Authors:** Amir Mousavi

## Abstract

Degenerative conditions of the lumbar spine, such as neural foraminal narrowing, subarticular stenosis, and spinal canal stenosis, are common and can lead to chronic pain and disability. Detecting and assessing the severity of these conditions is a key task in radiology, often requiring considerable expertise. In this work, we aim to develop automated models that aid radiologists in diagnosing these conditions using lumbar spine MR images. We utilize state-of-the-art deep learning methods for both detection and classification tasks, focusing on severity classification across five intervertebral disc levels and three different imaging modalities. The dataset and methodology used in this study are part of the RSNA 2024 Lumbar Spine Degenerative Classification competition. Our approach involves a two-step process, combining object detection with YOLO v8 and severity classification using a DeepScoreNet architecture. Through careful cross-validation and model evaluation, we demonstrate the potential of these models to improve diagnostic accuracy and efficiency. The DeepScoreNet model, introduced in this article, comprises 2,787,651 parameters and achieved accuracies of 81% for score(severity) prediction in cases of Neural Foraminal Narrowing, 89% for Spinal Canal Stenosis, and 74% for Subarticular Stenosis.

The code for this approach is available in the GitHub repository.(https://github.com/liamirpy/Lumbar-Spine-Degenerative-Classification)

## I. Introduction

Lumbar spinal stenosis (LSS) is a serious condition that affects over 200,000 adults in the U.S. and can greatly impact quality of life. Most individuals with LSS experience lower back pain, which often prompts them to seek medical care. According to the World Health Organization, low back pain is the leading cause of disability worldwide, affecting 619 million people in 2020. [1] Most people experience low back pain at some point in their lives, with its frequency increasing with age. Pain and restricted mobility are often symptoms of spondylosis, a set of degenerative spine conditions that include intervertebral disc degeneration and narrowing of the spinal canal (spinal stenosis), subarticular recesses, or neural foramina, often causing compression or irritation of nerves in the lower back. Proper diagnosis and grading of these conditions help guide treatment and potential surgery to alleviate pain and improve patients’ quality of life [1].

Magnetic resonance imaging (MRI) provides a detailed view of the lumbar spine, including the vertebrae, discs, and nerves, enabling radiologists to assess the presence and severity of degenerative spine conditions. MRI is essential for accurately evaluating the central canal, lateral recesses, and neural foramina, with the extent of stenosis in each area guiding treatment decisions. However, providing detailed reports can be repetitive and time-consuming, and the grading systems for LSS lack standardization [2].

Several deep learning (DL) systems have been developed to assist in the interpretation of advanced imaging, such as knee MRI and CT angiography for cerebral aneurysms. Prior DL work in lumbar spine MRI has shown potential, particularly through convolutional neural networks (CNNs), which can automatically learn image features to perform classification [3]. Automated grading of LSS using CNNs may improve the consistency, accuracy, and objectivity of assessments. DL applications in lumbar spine MRI have primarily focused on vertebra counting or disc degeneration grading [4]. In 2017, Jamaludin et al. introduced SpineNet, a multitask architecture for classifying lumbar central canal stenosis and other spinal conditions, using binary classification (stenosis present or absent) based solely on sagittal MRI [5]. In 2018, Lu et al. developed Deep Spine, a DL algorithm to grade LSS at the central canal and neural foramina, relying on weakly supervised natural language processing from radiology reports without a predefined grading system [6].

RSNA, in partnership with the American Society of Neuroradiology (ASNR), has organized a competition to explore whether artificial intelligence can be used to aid in detecting and classifying degenerative spine conditions using lumbar spine MRI. The competition focuses on five conditions: Left Neural Foraminal Narrowing, Right Neural Foraminal Narrowing, Left Subarticular Stenosis, Right Subarticular Stenosis, and Spinal Canal Stenosis. Severity scores (Normal/Mild, Moderate, or Severe) for these conditions are provided across the intervertebral disc levels L1/L2, L2/L3, L3/L4, L4/L5, and L5/S1. This dataset, sourced from eight sites on five continents, aims to improve the standardized classification of degenerative lumbar spine conditions and enable the development of tools to automate accurate and rapid disease classification [7].

To date, no DL model has been developed to assess stenosis across all three regions of interest (ROIs) in the lumbar spine. A model capable of this could provide radiologists with a reliable diagnostic tool for LSS. In this study, a DL model was created to automatically detect and classify stenosis in the central canal, lateral recess, and neural foramina using axial and sagittal MRI sequences.

To aid in this process, we propose an automated deep learning-based approach for detecting and classifying degenerative lumbar spine conditions. This study is based on the RSNA 2024 Lumbar Spine Degenerative Classification competition dataset, which includes MR images from three different modalities—Sagittal T1, Sagittal T2, and Axial T2—corresponding to specific conditions at various intervertebral disc levels (L1/L2, L2/L3, L3/L4, L4/L5, L5/S1). The goal is to classify these conditions into severity categories of *Normal/Mild, Moderate*, or *Severe*.

## II. Data Description

### A. Training Data

The training dataset contains the labels necessary for model training. Each row corresponds to a unique study of a patient, capturing relevant details about their spinal condition. The key columns include:

- **study id**: A unique identifier for each study. Each study may consist of multiple series of images, representing different orientations or slices of the lumbar spine.
- **condition [level]**: The target labels for various conditions affecting different vertebral levels, such as `spinal canal stenosis l1 l2`. These labels are categorized into severity levels: Normal/Mild, Moderate, or Severe. Some entries may contain incomplete labels.

This dataset is integral for developing machine learning models aimed at classifying lumbar spine conditions using image-based features and associated metadata.

### B. Label Coordinates

The label coordinates dataset provides detailed information about specific regions in the MRI scans corresponding to labeled conditions. It includes the following columns:

- **study id**: The unique identifier for each study, consistent with the training dataset.
- **series id**: An identifier for the particular image series within a study.
- **instance number**: The index of the image slice within a 3D stack, indicating its position in the sequence.
- **condition**: The medical condition being labeled, such as `spinal_canal_stenosis`, `neural _foraminal_narrowing`, or `subarticular_stenosis`. For some conditions, such as `neural foraminal_narrowing`, both left and right sides of the spine are considered.
- **level**: The vertebrae level associated with the condition, such as `l3 l4`.
- **x/y**: The coordinates specifying the center of the region in the image corresponding to the labeled condition.

These coordinates allow for the localization of pathologies in the MRI scans, enabling the creation of models that incorporate spatial information from the images.

### C. Imagery Data

The MRI scans are stored in the Digital Imaging and Communications in Medicine (DICOM) format, which is standard for medical imaging. The file structure is organized as follows:

~~~
[train/test] images/[study id]/
[series id]/[instance number].dcm
~~~

Each `study_id` contains multiple `series_id`s, representing different orientations or imaging sequences of the lumbar spine. Within each series, the `instance_number` identifies individual slices of the 3D MRI stack. The DICOM files contain both image data and metadata crucial for interpreting the scans.

### D. Series Descriptions

The series descriptions metadata provides information about the different MRI scan series. The key columns include:

- **study id**: The unique identifier for each study.
- **series id**: The identifier for the image series within the study.
- **series description**: A description of the scan’s orientation or protocol used during acquisition, such as `sagittal` or `axial`.

This metadata is useful for distinguishing between different imaging orientations and protocols, aiding in the analysis of the scan data.

## III. Methodology

Our approach consists of two main steps: object detection and classification. Each part is described in detail below.

### A. Data Preprocessing and Cleaning

The first step involves preprocessing and cleaning the data. The train label coordinates.csv file provides the x and y coordinates of regions in the MR images associated with different conditions. These coordinates were used to crop the regions of interest (ROI) from the full images. Additional metadata such as the series descriptions and severity scores were merged with the coordinates to create a single comprehensive dataset (saved as dataset description.csv) for further use.

After cleaning, the data was split into three separate CSV files for each of the conditions: **Neural Foraminal Narrowing, Spinal Canal Stenosis**, and **Subarticular Stenosis**, which were used to train the respective models. The corresponding conditions on the left and right sides were grouped together.

### B. Object Detection with YOLO v8

The first stage of our pipeline involves detecting the region of interest (ROI) associated with each condition. For this purpose, we employed the YOLO v8 object detection model, which is known for its speed and accuracy. Separate YOLO models were trained for each condition, focusing on the following:

- **Neural Foraminal Narrowing:** A total of 10 classes (2 sides * 5 intervertebral levels).
- **Spinal Canal Stenosis:** 5 classes (1 condition * 5 intervertebral levels).
- **Subarticular Stenosis:** 10 classes (2 sides * 5 intervertebral levels).

The training process involved converting the MR images from DICOM format to PNG with pixel normalization. The ground truth labels were prepared in YOLO format, where each bounding box is represented as: `class_id center_x center_y width height`.

For model evaluation, we implemented 5-fold cross-validation, ensuring that each fold maintained the same distribution of condition and intervertebral levels.

### C. Classification with DeepScoreNet

After detecting the ROIs, the next step was to classify the severity of the detected condition. For this, we employed a DeepScoreNet model pre-trained on the ImageNet dataset, fine-tuning it to classify the severity of degenerative spine conditions. Separate classifiers were trained for each condition:

- **Neural Foraminal Narrowing:** Classified into Normal/Mild, Moderate, or Severe.
- **Spinal Canal Stenosis:** Classified into Normal/Mild, Moderate, or Severe.
- **Subarticular Stenosis:** Classified into Normal/Mild, Moderate, or Severe.

#### 1) Handling Class Imbalance

The dataset exhibited significant class imbalance, with some severity levels having far fewer samples. To address this, we used a combination of data augmentation and a custom loss function based on class distribution. The data augmentation strategy involved oversampling the minority classes to balance the dataset as follows:

- The first minority class was augmented until it reached one-third of the size of the majority class.
- The second minority class was augmented until it reached half of the size of the majority class.

This approach helped to mitigate overfitting while maintaining a balanced training set.

### D. K-Fold Cross-Validation

For both the detection and classification tasks, we used 5-fold cross-validation. Each fold was split based on condition and intervertebral level, ensuring that the distribution of conditions was consistent across all folds. After splitting, the training data was augmented and passed through the YOLO and DeepScoreNet models for training.

## IV. Results

### A. The result of Condition Detection (YOLOv3)

**Fig. 1.**
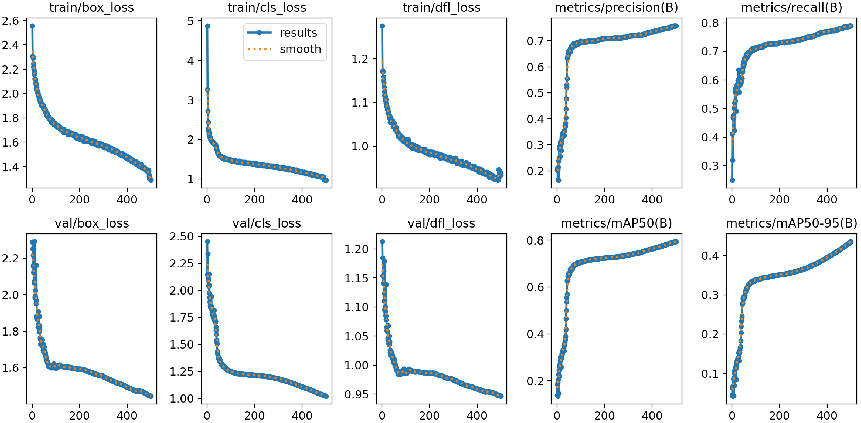
Neural Foraminal Narrowing fold 0

**Fig. 2.**
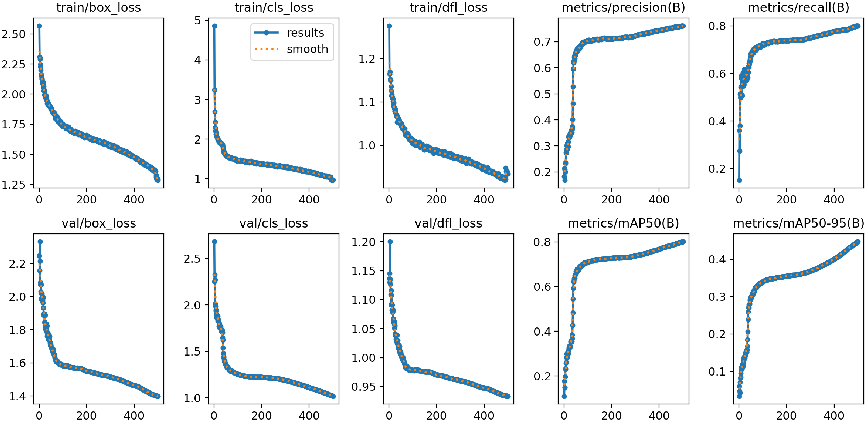
Neural Foraminal Narrowing fold 1

**Fig. 3.**
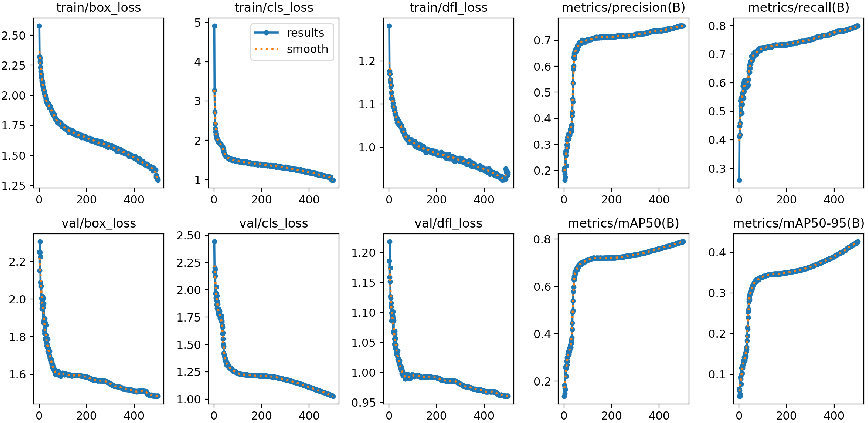
Neural Foraminal Narrowing fold 2

**Fig. 4.**
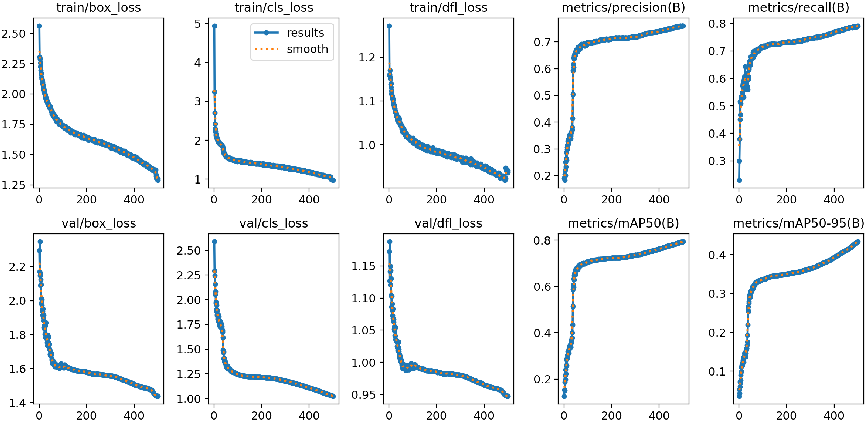
Neural Foraminal Narrowing fold 3

**Fig. 5.**
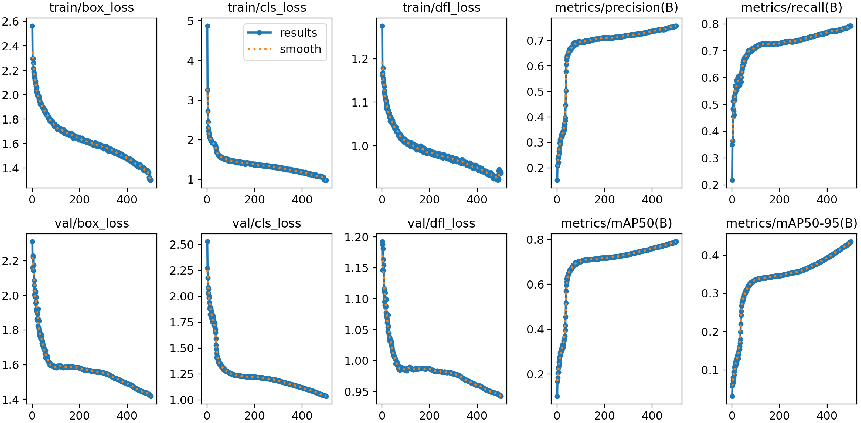
Neural Foraminal Narrowing fold 4

#### 1. Neural Foraminal Narrowing

**Fig. 6.**
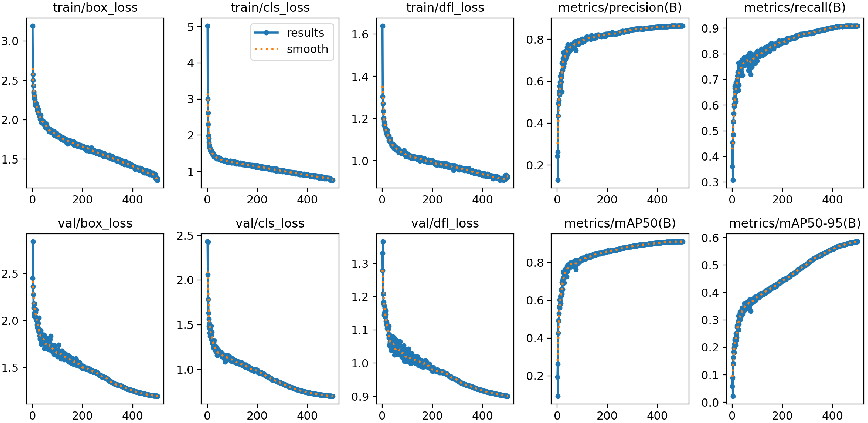
Spinal Canal Stenosis fold 0

**Fig. 7.**
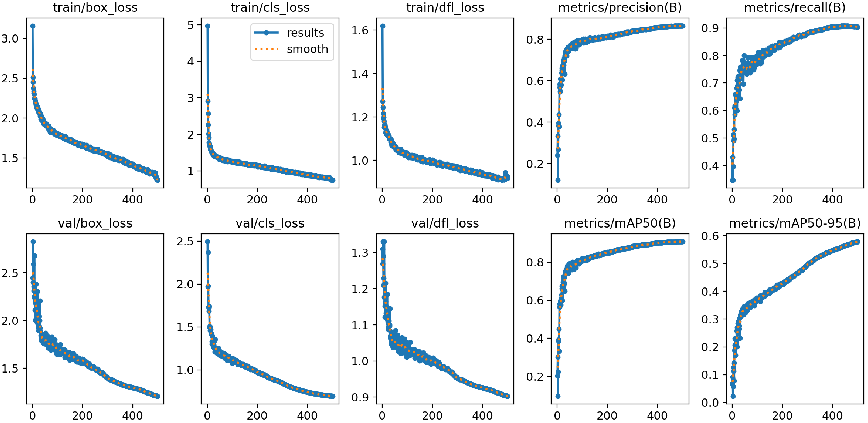
Spinal Canal Stenosis fold 1

**Fig. 8.**
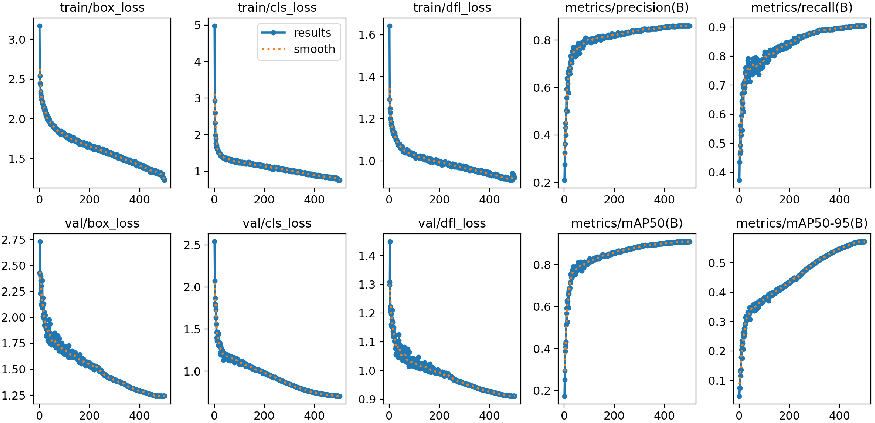
Spinal Canal Stenosis fold 2

**Fig. 9.**
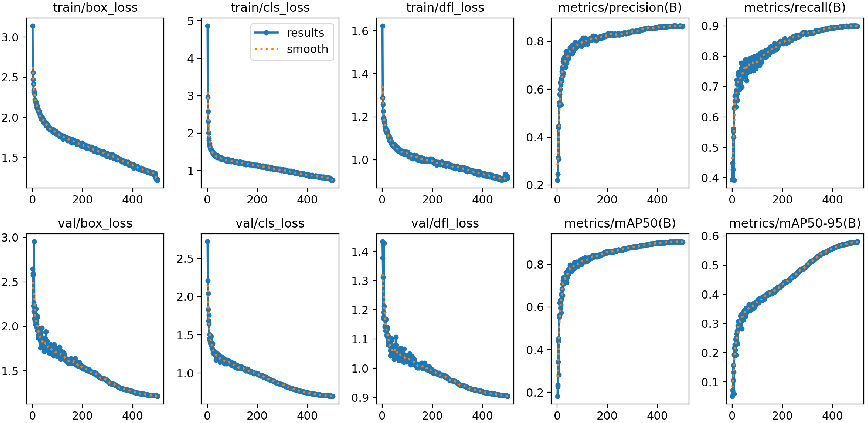
Spinal Canal Stenosis fold 3

**Fig. 10.**
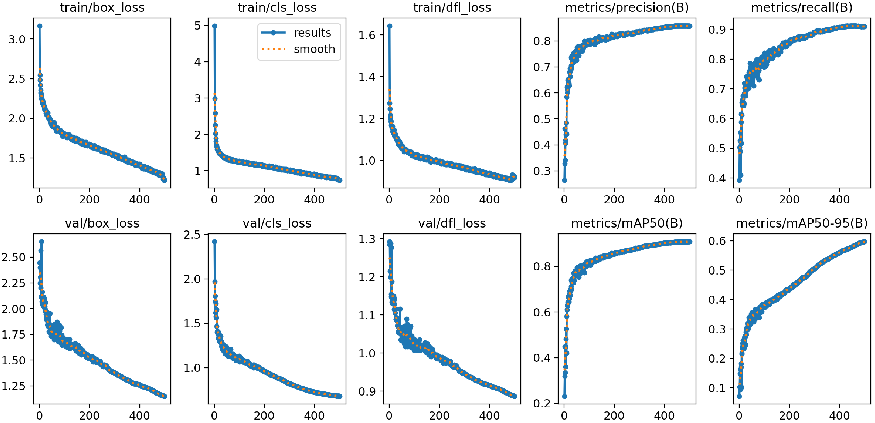
Spinal Canal Stenosis fold 4

#### 2. Spinal Canal Stenosis

**Fig. 11.**
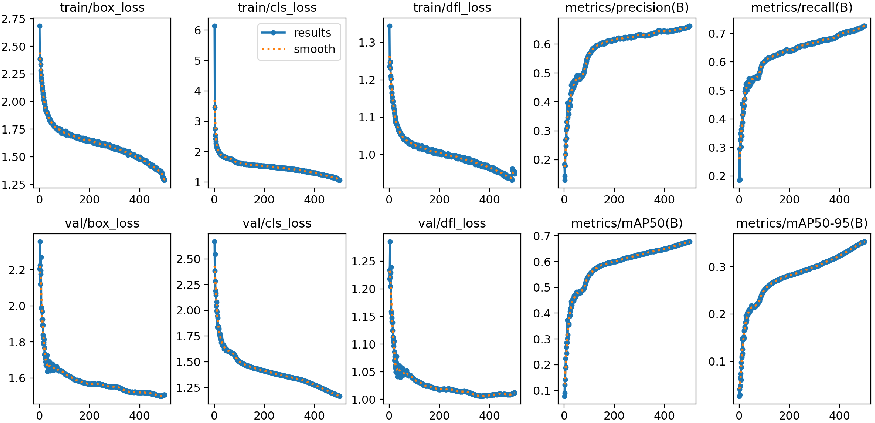
Subarticular Stenosis fold 0

**Fig. 12.**
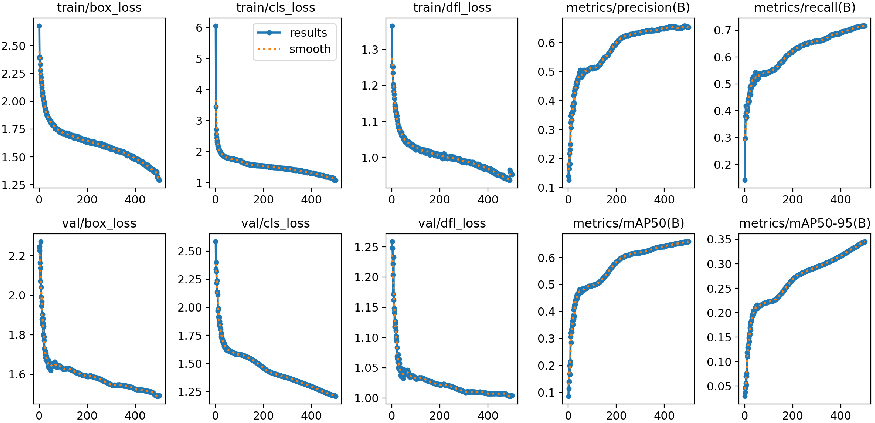
Subarticular Stenosis fold 1

**Fig. 13.**
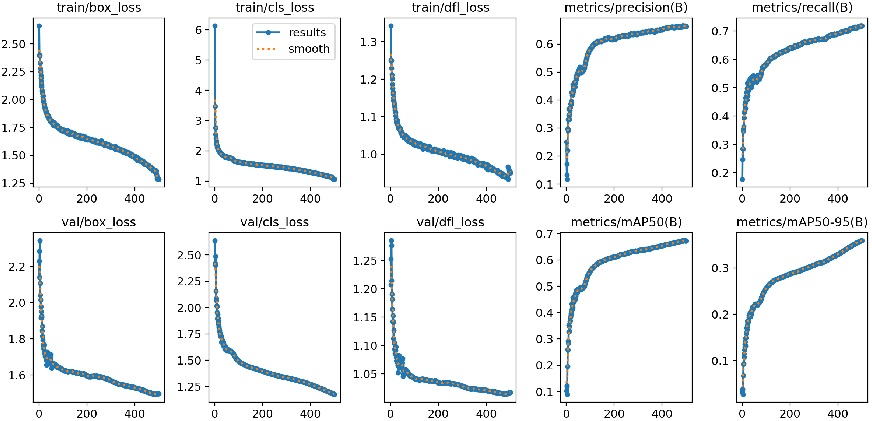
Subarticular Stenosis fold 2

**Fig. 14.**
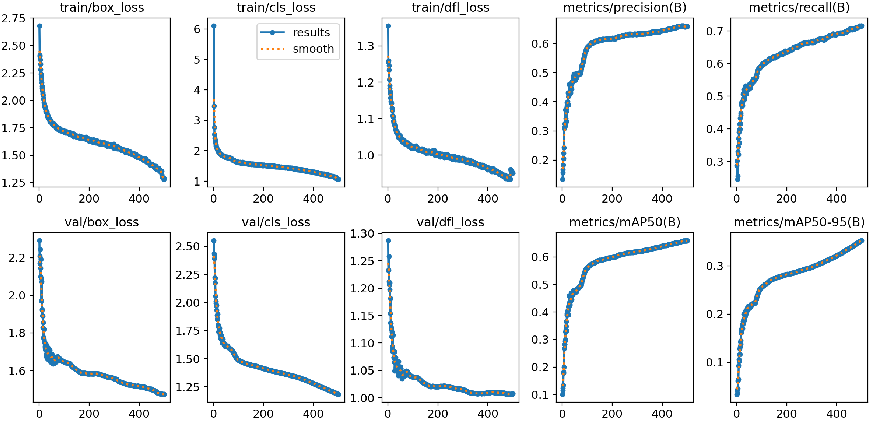
Subarticular Stenosis fold 3

**Fig. 15.**
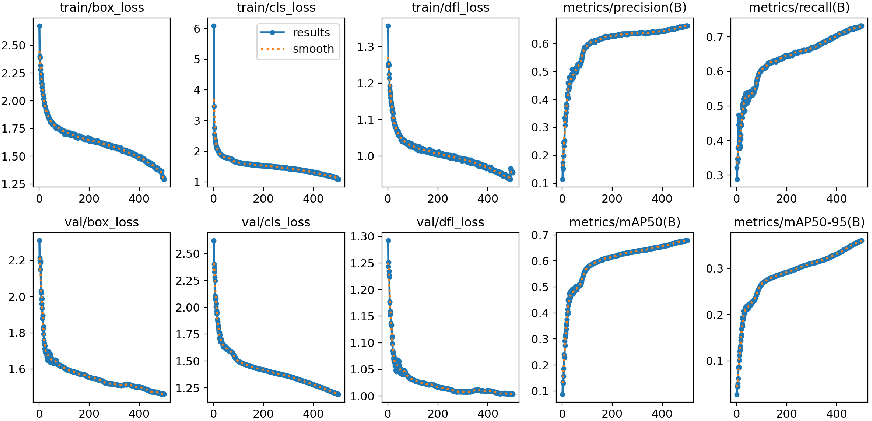
Subarticular Stenosis fold 4

#### 3. Subarticular Stenosis

### B. The result of Score(severity) prediction (DeepScoreNet)

**TABLE I.** Cross-validation results for Neural Foraminal Narrowing.

| Fold | Accuracy | Precision | Recall | F1 Score |
| --- | --- | --- | --- | --- |
| Fold 0 | 0.7801 | 0.8346 | 0.7801 | 0.7984 |
| Fold 1 | 0.7852 | 0.8385 | 0.7852 | 0.8018 |
| Fold 2 | **0.8121** | 0.8208 | **0.8121** | **0.8161** |
| Fold 3 | 0.7976 | 0.8292 | 0.7976 | 0.8096 |
| Fold 4 | 0.7978 | 0.8310 | 0.7978 | 0.8105 |

1. *Neural Foraminal Narrowing:* Fold 2 achieved the highest accuracy (0.8121) and F1 score (0.8161), making it the best-performing fold. This fold was selected for further evaluation and deployment.
2. *Spinal Canal Stenosis:* Fold 3 yielded the highest accuracy (0.8995) and F1 score (0.9051), demonstrating the model’s ability to detect Spinal Canal Stenosis with high reliability.
3. *Subarticular Stenosis:* Fold 2 achieved the highest performance for Subarticular Stenosis, with an accuracy of 0.7931 and an F1 score of 0.8034.

**TABLE II.** Cross-validation results for Spinal Canal Stenosis.

| Fold | Accuracy | Precision | Recall | F1 Score |
| --- | --- | --- | --- | --- |
| Fold 0 | 0.8954 | 0.9025 | 0.8954 | 0.9008 |
| Fold 1 | 0.8898 | 0.8973 | 0.8898 | 0.8940 |
| Fold 2 | 0.8935 | 0.8984 | 0.8935 | 0.8955 |
| Fold 3 | **0.8995** | 0.9017 | **0.8995** | **0.9051** |
| Fold 4 | 0.8973 | 0.9019 | 0.8973 | 0.9021 |

**TABLE III.** Cross-validation results for Subarticular Stenosis.

| Fold | Accuracy | Precision | Recall | F1 Score |
| --- | --- | --- | --- | --- |
| Fold 0 | 0.7791 | 0.7824 | 0.7791 | 0.7807 |
| Fold 1 | 0.7902 | 0.8031 | 0.7902 | 0.7951 |
| Fold 2 | **0.7931** | 0.8087 | **0.7931** | **0.8034** |
| Fold 3 | 0.7908 | 0.8073 | 0.7908 | 0.8021 |
| Fold 4 | 0.7919 | 0.8092 | 0.7919 | 0.8055 |

## V. Conclusion

The proposed two-step deep learning pipeline for lumbar spine degenerative classification, using YOLO for condition detection and VGG-19 for severity classification, showed promising results. The imbalance in severity classes was effectively managed using data augmentation and a custom loss function.

**The important aspect of this approach is that it performs exceptionally well in the score prediction component. However, the condition detection does not function satisfactorily. The aim of publishing this article is to encourage other researchers to focus more on the detection aspect to enhance performance. This improvement is crucial because the score detection phase follows the condition detection in a sequential manner; therefore, enhancing the detection component will lead to an overall improvement in results. In the score prediction segment, the model was developed by myself, and we successfully reduced the number of trainable parameters while achieving excellent performance in score detection**.

## Data Availability

The dataset and methodology used in this study are part of the RSNA 2024 Lumbar SpineDegenerative Classification competition.

https://www.kaggle.com/competitions/rsna-2024-lumbar-spine-degenerative-classification

---

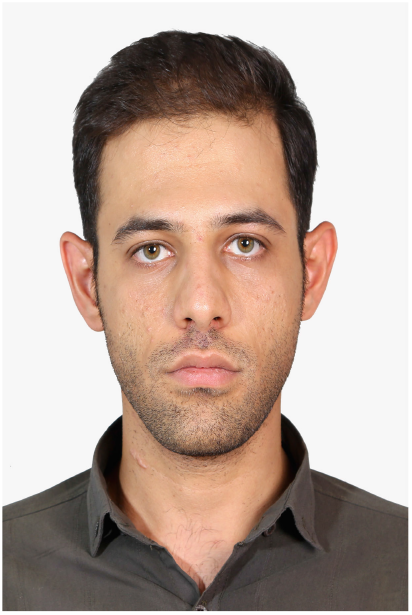

Sayed Amir Mousavi Mobarakeh holds a B.Sc. in Electrical Engineering from Isfahan University (2019) and an M.Sc. from Shiraz University of Technology (2023). Currently pursuing a Ph.D. in Neuroscience at UPJV, France, his research integrates medical imaging, machine learning, and innovative architectural design for diverse medical applications. During his master’s, he focused on COVID-19 CT image segmentation, showcasing dedication to addressing critical healthcare challenges. Mousavi Mobarakeh’s academic trajectory highlights his commitment to advancing neuroscience and medical engineering, positioning him as a promising researcher at the intersection of technology and healthcare.

